# Features and Functions of Systemic and Mucosal Humoral Immunity Among SARS-CoV-2 Convalescent Individuals

**DOI:** 10.1101/2020.08.05.20168971

**Authors:** Savannah E. Butler, Andrew R. Crowley, Harini Natarajan, Shiwei Xu, Joshua A. Weiner, Jiwon Lee, Wendy Wieland-Alter, Ruth I. Connor, Peter F. Wright, Margaret E. Ackerman

**Author notes:** Contributed equally (Savannah E. Butler, Andrew R. Crowley, Harini Natarajan). Corresponding Authors Peter F. Wright 1 Medical Center Drive Lebanon, NH 03756 (ph), Margaret E. Ackerman 14 Engineering Drive Hanover, NH 03755, (ph).

## Abstract

Understanding humoral immune responses to SARS-CoV-2 infection will play a critical role in the development of vaccines and antibody-based interventions. We report systemic and mucosal antibody responses in convalescent individuals who experienced varying disease severity. Robust antibody responses to diverse SARS-CoV-2 antigens and evidence of elevated responses to endemic CoV were observed among convalescent donors. SARS-CoV-2-specific IgA and IgG responses were often negatively correlated, particularly in mucosal samples, suggesting subject-intrinsic biases in isotype switching. Assessment of antibody-mediated effector functions revealed an inverse correlation between systemic and mucosal neutralization activity and site-dependent differences in the isotype of neutralizing antibodies. Serum neutralization correlated with systemic anti-SARS-CoV-2 IgG and IgM response magnitude, while mucosal neutralization was associated with nasal SARS-CoV-2-specific IgA. These findings begin to map how diverse Ab characteristics relate to Ab functions and outcomes of infection, informing public health assessment strategies and vaccine development efforts.

## Introduction

Since its emergence in late 2019 in China’s Hubei province, SARS-CoV-2, the human coronavirus (CoV) causing COVID-19, has spread rapidly. Formal designation as a pandemic followed in March of 2020, making understanding the health implications of infection and the development of effective interventions a global priority. To this end, studies of endemic and other pathogenic CoV strains and evaluation of humoral immune responses induced by SARS-CoV-2 infection in humans have contributed to our understanding of how antibodies (Abs) might provide protection from infection or severe disease, or alternatively, contribute to disease pathology.

Due to the critical role of the spike (S) protein in viral entry, Abs targeting S, particularly in the receptor binding domain (RBD), have been shown to neutralize the infectivity of CoVs (Chan et al., 2009; Sui et al., 2004; ter Meulen et al., 2006; Traggiai et al., 2004; Zhu et al., 2007) including SARS-CoV-2 (Chen et al., 2020; Wang et al., 2020; Ye et al., 2020). Studies of monoclonal Abs isolated from SARS-CoV-2- infected individuals have shown potent anti-viral effects *in vitro* and protection from viral challenge in mouse and nonhuman primate models (Cao et al., 2020; Hassan et al., 2020; Imai et al., 2020; Ju et al., 2020; Pinto et al., 2020; Rogers et al., 2020; Salazar et al., 2020; Shi et al., 2020; Wec et al., 2020; Wu et al., 2020; Yuan et al., 2020). Similarly, polyclonal Ab responses observed in convalescent and vaccinated macaques have demonstrated promising protection profiles in challenge experiments (Chandrashekar et al., 2020; Yu et al., 2020). Collectively, these studies establish firm proof of principle for Ab-mediated protection, motivating passive transfer of polyclonal serum Abs from recovered individuals as a clinical intervention (Duan et al., 2020; Shen et al., 2020; Xu et al., 2020; Zeng et al., 2020; Zhang et al., 2020). These efforts are challenged by the diversity of serum responses observed among convalescent donors (Klein et al., 2020), and variability in the symptoms and sequelae of treated individuals (Guan et al., 2020). Moreover, some CoV-specific Abs have been reported to contribute to disease pathology: while the mechanism whereby SARS-CoV-2 sometimes triggers cytokine storm remain to be fully elucidated, Ab-Dependent Enhancement (ADE) of infection or disease has been implicated in the pathogenesis of SARS-CoV-2 and other CoV (Cong et al., 2018; Hoeppel et al., 2020; Jaume et al., 2011; Liu et al., 2019; Vennema et al., 1990; Yang et al., 2005; Yip et al., 2014).

Other challenging aspects of understanding humoral immunity to SARS-CoV-2 are that the highest Ab titers and most potent neutralizing Ab responses have been observed in individuals with severe infection (Klein et al., 2020; Long et al., 2020b), while infected subjects with mild symptoms may not seroconvert (Gallais et al.; Le Bert et al., 2020; Sekine et al., 2020). Further, neutralizing Ab responses have been observed to wane quickly (Long et al., 2020b; Seow et al.), suggesting the full spectrum of anti-viral functions of Abs and T cells may be needed to contribute to protection from infection or disease. As a result, there is concern that neither vaccines nor prior infection will be highly effective, that any protective effects may have limited durability, and that the most vulnerable individuals, such as the immunocompromised, elderly, and those with co-morbidities, may have an inadequate level of protection. Cumulatively, many questions remain about the features and functions of systemic and mucosal SARS-CoV-2-reactive Abs present after infection and how they may contribute to viral clearance or inflammatory pathology.

To characterize the systemic and mucosal humoral immune responses to SARS-CoV-2 and better understand the dynamic between Ab features and functions within each compartment, we turned to systems serology (Pittala et al., 2019). This technique utilizes high-throughput, multidimensional biophysical profiling of Ab response features, cell-based assays of Ab functions, and machine learning as a means to discover mechanistically meaningful signatures of Ab-mediated protection and activity beyond response magnitude (Barouch et al., 2015). We assessed antigen binding, CoV-specific Ab isotypes and subclasses, and binding to FcγR and FcαR. These biophysical Ab response features were complemented by functional analysis of neutralization and Ab-mediated effector functions with the goal of defining the sites and characteristics of functionally potent humoral immune responses.

## Results

### Systemic and Mucosal SARS CoV-2 Specific Ab Response Features

Serum, nasal wash, and stool samples were collected approximately one month (range: 19-67 days, mean: 40 days) after initial clinical presentation from 20 subjects (age range: 18-77, mean: 53 years) who tested positive for SARS CoV-2 by qPCR, and from 15 SARS-CoV-2 naïve subjects (age range: 22-66, mean: 40 years). Ab responses to SARS-CoV-2 were evaluated using an Fc array (Brown et al., 2017; Brown et al., 2018) to characterize isotypes, subclasses, and Fc Receptor (FcR) binding across Abs specific to a panel of SARS-CoV-2 antigens. This panel included spike protein in trimeric, subdomain (i.e. S1, S2), and receptor binding domain (RBD) forms, nucleocapsid (N) protein, and the fusion peptide. S proteins from seven other endemic CoV strains, those associated with prior outbreaks (SARS-CoV-1 and MERS), and a closely related bat coronavirus, WIV1 (77.1% S amino acid identity) (Uddin et al., 2020), plus two non-CoV control antigens (influenza HA and herpes simplex virus gE), were also evaluated. Our results demonstrate SARS-CoV-2-specific Ab responses in COVID-19 convalescent but not naïve donor sera and nasal wash (**Fig. 1A**). Convalescent donor samples showed strong cross-reactive responses to the closely related bat CoV, WIV1. Elevated levels of OC43 (29.7% S amino acid identity) S-specific IgG and IgA Abs were also observed (unpaired two-tailed t-test with Welch’s correction, p < 0.0001, p = 0.011 respectively). This apparent boosting of responses to endemic CoV was observed more broadly among IgG1 and IgG3 subclass responses (**Supplemental Fig. 1**). Ab responses in serum and nasal wash samples were further examined by measuring levels of other Ab isotypes, subclasses, and by defining binding to diverse FcRs (**Fig. 1B, Supplemental Fig. 2-6**). SARS CoV-2-specific IgG1, IgG2, IgG3, IgA, IgM, and Abs able to ligate FcRs (FcαR, FcgR) were observed among samples from convalescent donors but not naïve subjects. In contrast to the robust responses apparent in serum and nasal samples, limited SARS CoV-2 specific Ig was detected in stool samples (**Supplemental Figure 7**), though like serum and nasal samples, responses toward OC43 appeared elevated in a fraction of convalescent donors. Systematic analysis of the magnitude and statistical confidence of differences in Ab present in convalescent versus naïve donors indicated elevated IgG, IgA, and IgM responses across diverse SARS CoV-2 antigen types as well as a number of endemic and related beta-CoV (**Supplemental Figure 8**).

**Figure 1.**
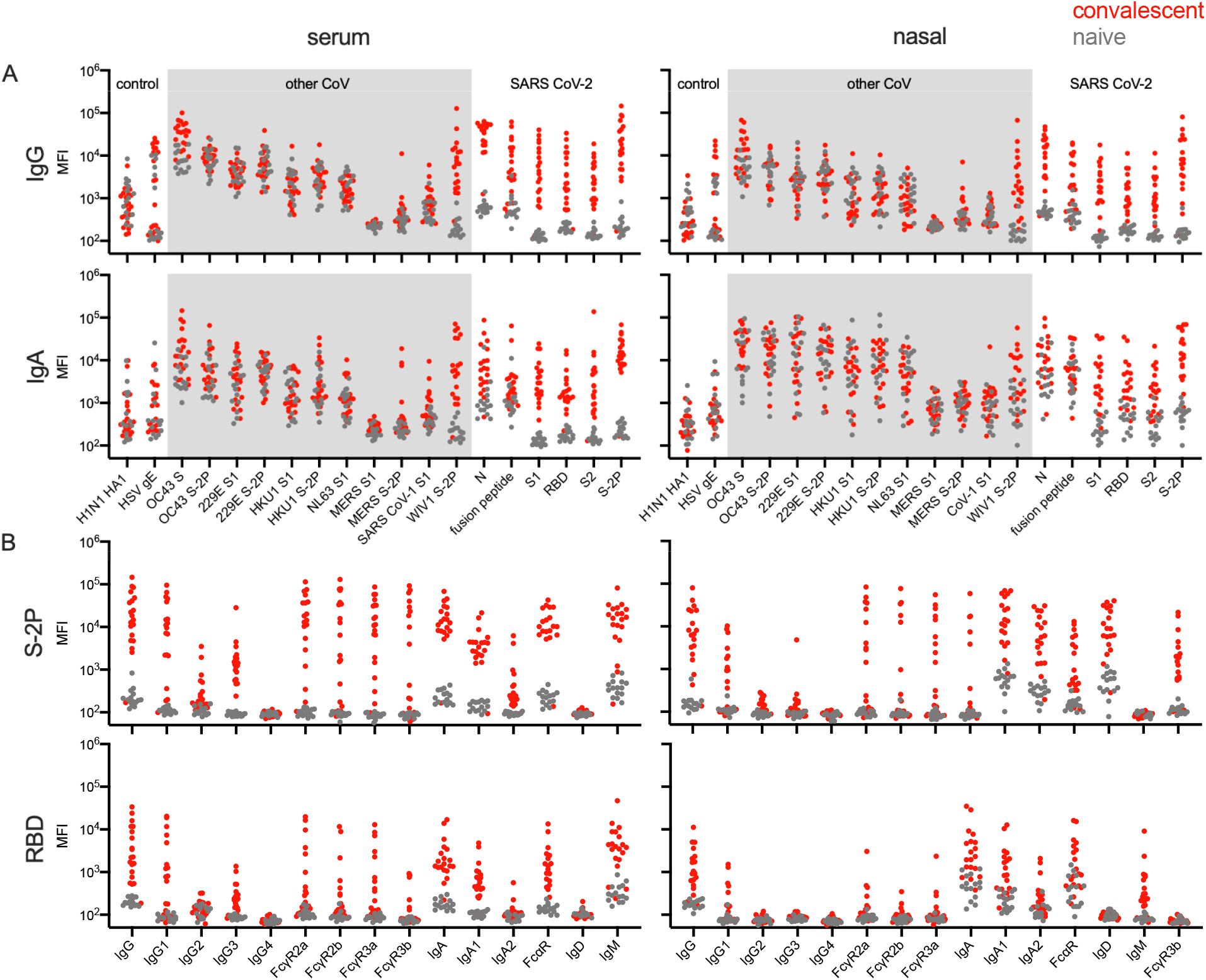
Systemic and mucosal Ab responses. **A**. Fc array characterization of IgG (top) and IgA (bottom) responses against a panel of SARS-CoV-2, other CoV, and control antigens in serum (left) and nasal wash (right) from convalescent (red) and naive (gray) donors. **B**. Abs to SARS CoV-2 S (S-2P) and receptor binding domain (RBD) across isotypes, subclasses, and for FcR binding as measured in serum (left) and nasal wash (right).

**Figure 2.**
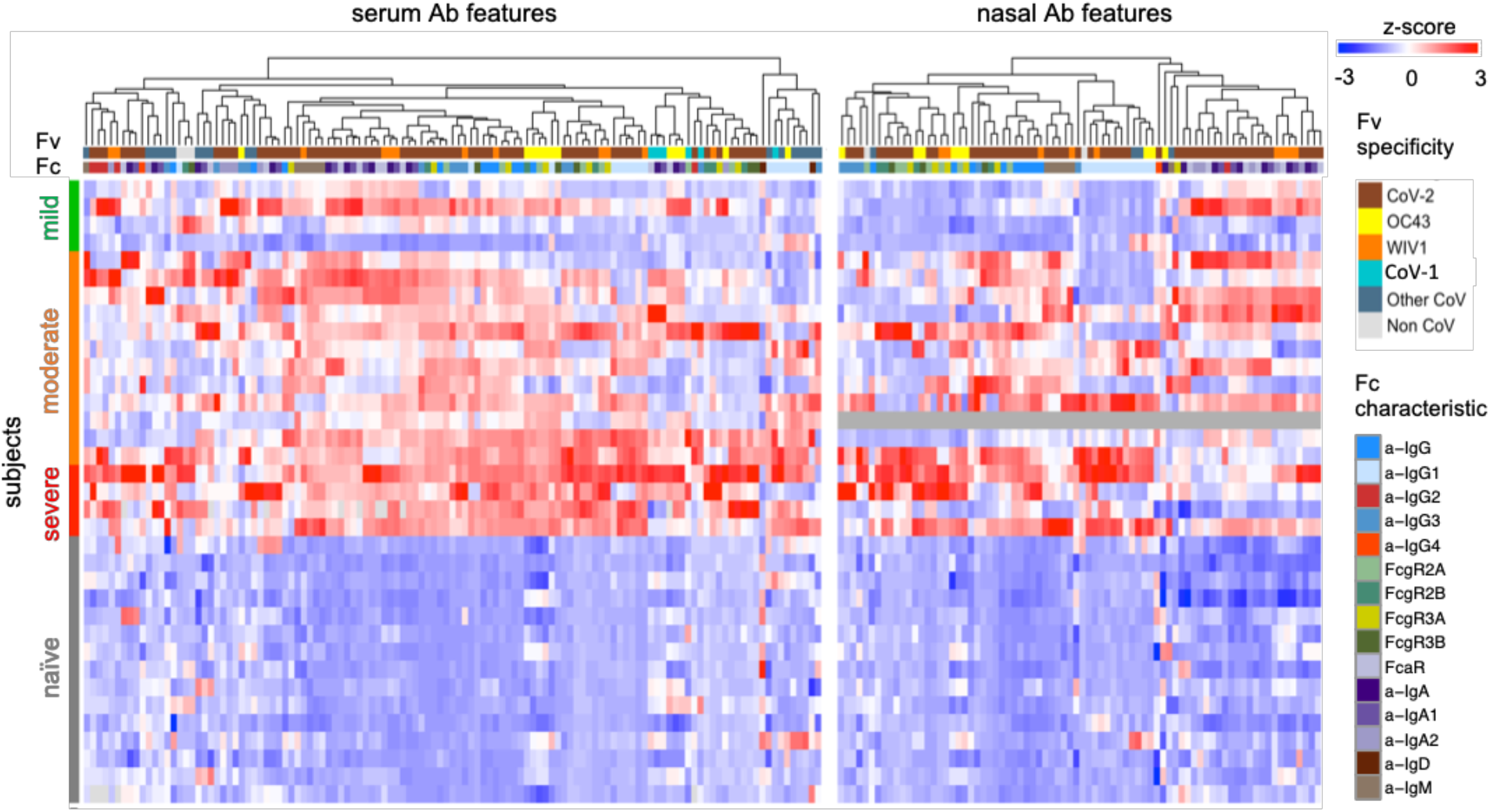
Relationships among subjects and Ab features in serum and nasal wash. Heatmap of filtered and hierarchically-clustered Fc array features in serum (left) and nasal wash (right) across subjects with varying infection or disease status. Responses are centered and scaled per feature and the scale range truncated at +/-3 SD. Antigen specificity (Fv) and Fc characteristics (Fc) are indicated in color bars.

To understand how aspects of the humoral response relate to each other, hierarchical clustering was performed on Ab features to define similarities among subjects and features (**Supplemental Figures 9-10**). When focusing on those features that were significantly increased among convalescent donors (unpaired two-tailed t-test with Welch’s correction, p < 0.05), elevated IgG responses were observed in both serum and nasal samples in convalescent donors who had experienced severe disease (**Fig. 2**). In contrast, elevated nasal IgA responses were apparent among donors with mild or moderate disease. When correlations between Ab types and specificities elicited among convalescent donors were plotted for serum and nasal samples, clear patterns emerged suggesting biases between IgA and IgG responses. For example, in serum, IgG1, IgG3 and FcγR-binding Ab responses were well correlated with each other across diverse specificities, as were IgA, IgA1, and IgA2 and FcαR-binding Abs (**Fig. 3A**). Correlations between these isotypes were more modest, though overall in serum, many response features were positively correlated with each other. In contrast, nasal responses showed clear evidence of a bias to favor either IgG or IgA, as striking inverse correlations were observed between these isotypes across diverse antigen specificities (**Fig. 3B**).

**Figure 3.**
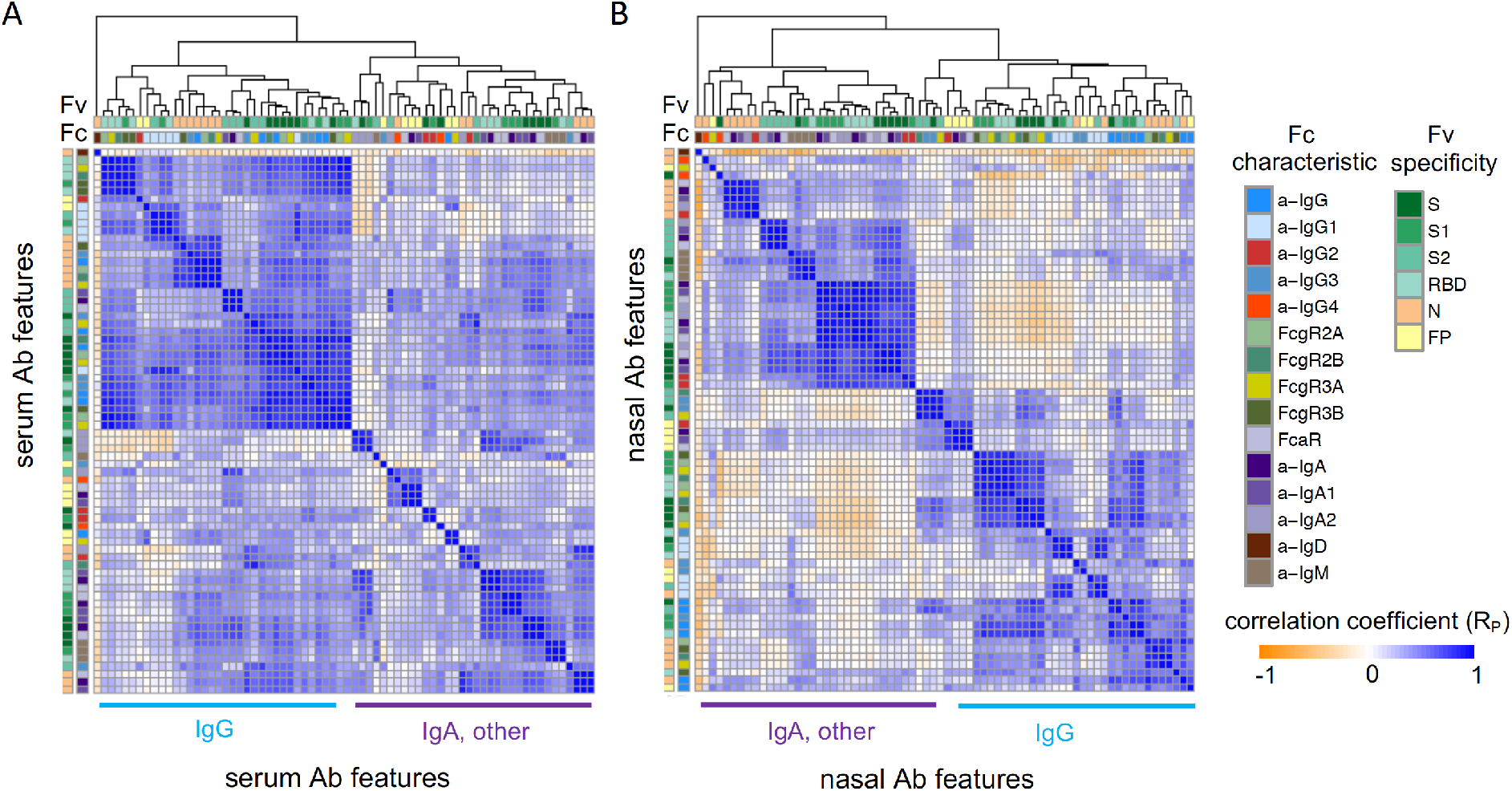
Correlations between Ab features within serum and nasal wash samples. **A-B**. Correlation matrices of relationships between Ab response features measured in serum (**A**) and nasal wash (**B**) samples from convalescent donors. Antigen specificity (Fv) and Fc characteristics (Fc) are indicated in color bars. Filtered CoV-2-specific Ab features are hierarchically clustered. Pearson correlation coefficients (Rp) are shown. Dominant isotype(s) of main branches of dendrograms are indicated.

Because mucosal and systemic responses can exhibit remarkable divergence (Wright et al., 2016), we determined correlations in the humoral immune response between these anatomical sites using sera and nasal samples from convalescent donors (**Fig. 4A**). IgG and IgA responses in serum were directly correlated with IgG and IgA responses, respectively, in nasal wash samples. However, no or inverse relationships were observed between the serum IgG and nasal IgA; between serum IgA and nasal IgG; or between different isotypes at different anatomic sites, as shown for a representative S antigen (**Fig. 4B**).

**Figure 4.**
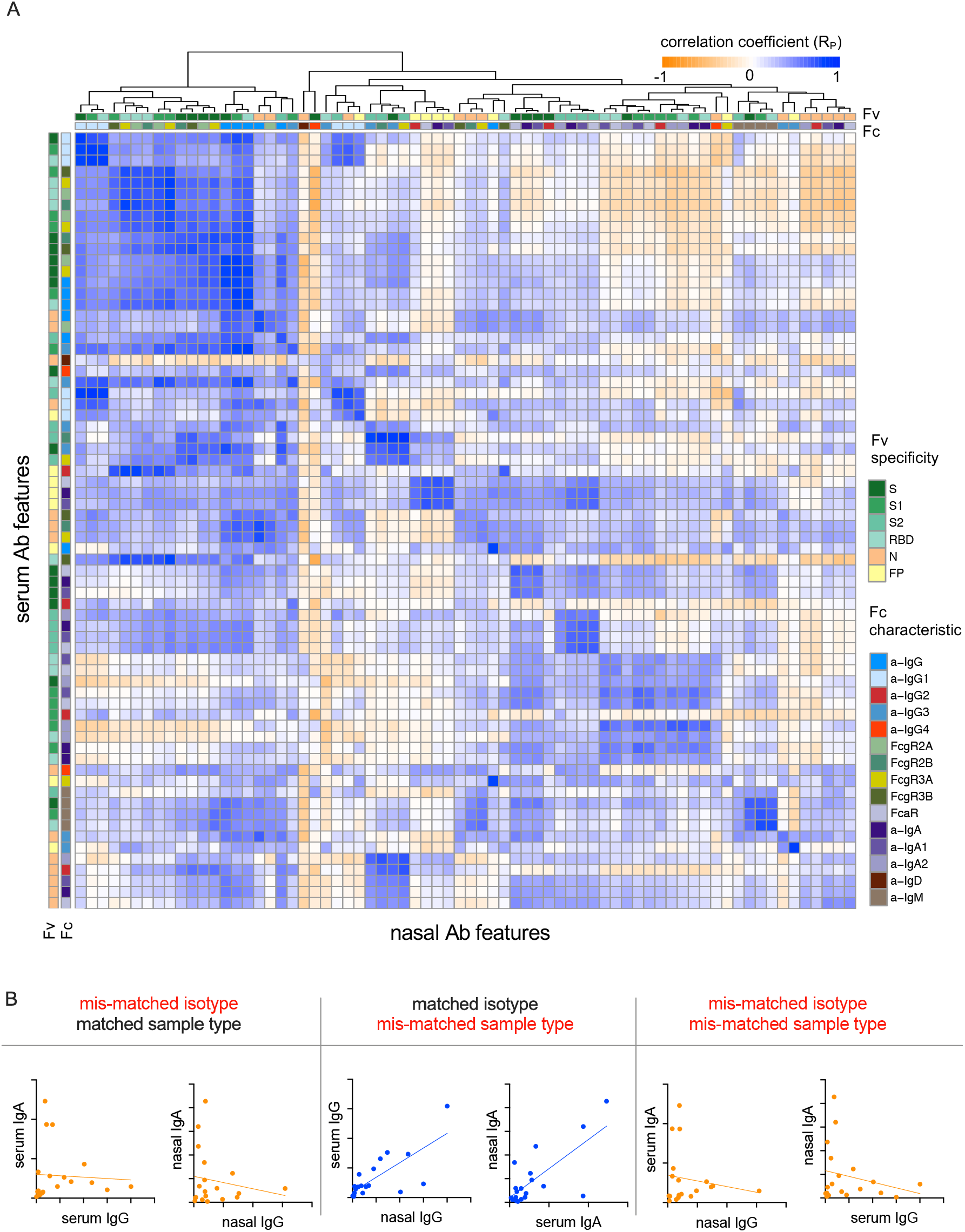
Correlation between Ab response in serum and nasal wash samples. **A**. Correlation matrix of relationships between hierarchically-clustered Ab response features measured in serum and nasal wash samples from convalescent donors. Antigen specificity (Fv) and Fc characteristics (Fc) are indicated in color bars. Pearson correlation coefficients (R_p_) are shown. **B**. Representative scatter plots of the correlative relationships between IgA and IgG anti-S1 responses in nasal and serum samples.

### Neutralization activity of systemic and mucosal Ab

Given evidence of robust humoral responses in systemic and mucosal samples, we next sought to determine neutralization potency in both serum and nasal wash samples using a luciferase-based SARS-CoV-2 pseudovirus assay (Letko et al., 2020). Consistent with other studies (Klein et al., 2020), elevated serum neutralization activity was observed for subjects who experienced severe, as compared to nonsevere, disease (unpaired two-tailed t-test with Welch’s correction, p = 0.034) (**Fig. 5a**). In contrast to observations in serum, nasal samples from subjects with severe disease showed little to no viral neutralization, whereas subjects with elevated mucosal neutralization activity tended to have experienced mild or moderate symptoms (**Fig. 5b**). Indeed, nasal and serum neutralization activities exhibited an inverse relationship (**Fig. 5c**). This observation further suggests the potential importance of the observed distinctions in Ab isotypes between mucosal and systemic Ab responses.

**Figure 5.**
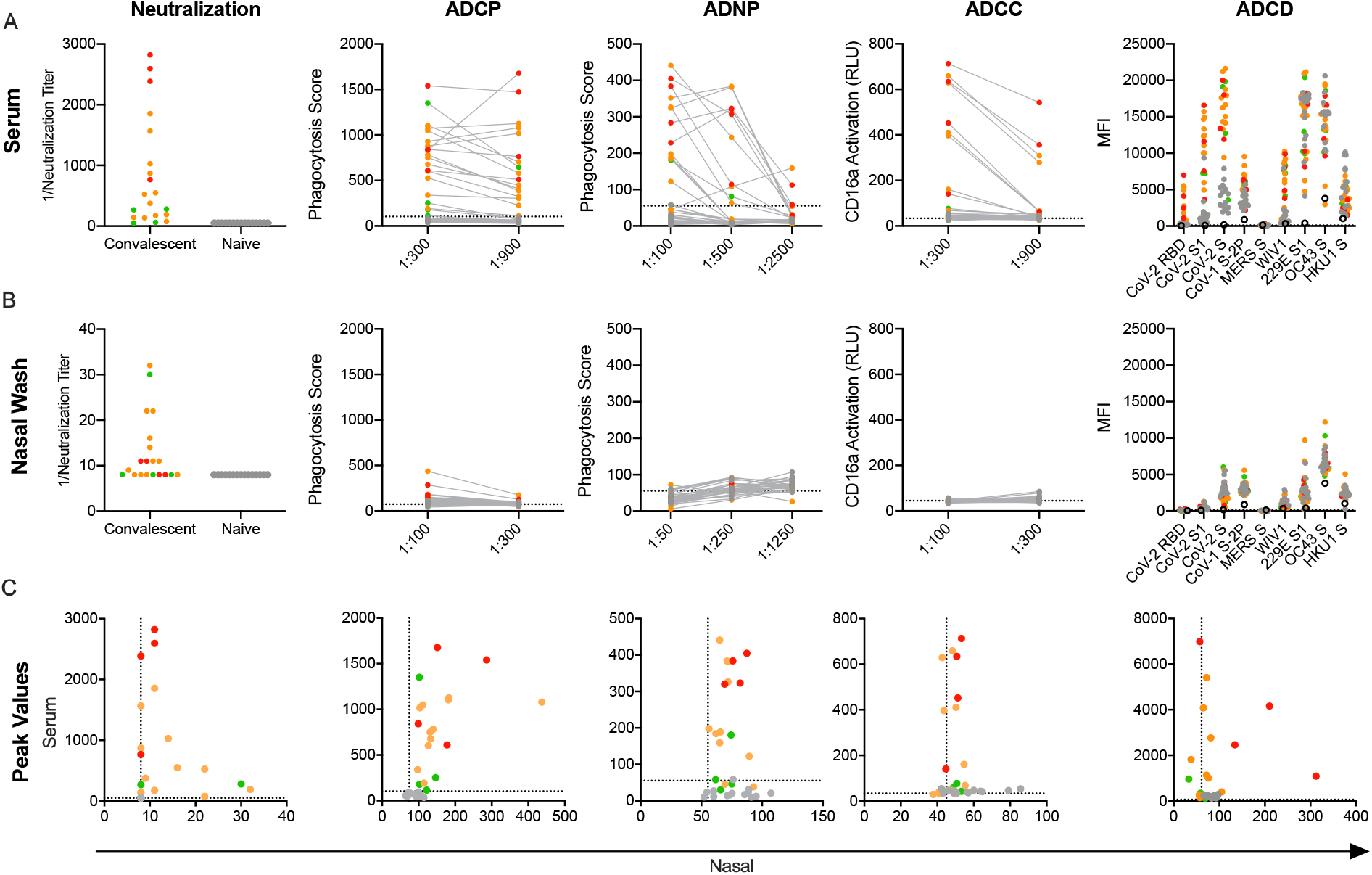
Mucosal and systemic Ab functions. **A,B**. Functional activity of serum (**A**) and (**B**) nasal wash subject samples in a panel of neutralization and effector function assays. **C**. Scatterplots of serum versus mucosal activities observed for each subject for viral neutralization and RBD-specific Ab effector functions. Titer is plotted for neutralization data and peak activity is plotted for effector functions. Infection status and disease severity is indicated in color. Limit of detection (neutralization) or values observed for no Ab controls (ADCP, ADNP, ADCC) are indicated with dotted lines. No Ab controls for ADCD are indicated with the hollow black circle.

### Effector functions of systemic and mucosal Ab

Beyond neutralization, however, little is known about the antiviral functions of systemic and mucosal Abs in COVID-19 convalescent donors. We sought to further characterize the antiviral activities of Abs in serum and nasal samples by evaluating their effector functions against CoV-2 RBD, including Ab-mediated phagocytosis, NK cell receptor ligation and complement activation. Our results demonstrate that serum from most convalescent subjects readily promoted phagocytosis mediated by monocyte (ADCP) and neutrophil (ADNP) effector cells (**Fig 5a**). While nasal wash samples were far less capable of driving functional activity, a number of subjects exhibited nasal Ab responses able to elicit phagocytosis in monocytes (**Fig. 5b**). Serum from these subjects also tended to generate a strong phagocytic response (**Fig 5c**). Across phagocytosis, NK cell FcgR3a receptor ligation (ADCC), and complement cascade protein C3b deposition (ADCD), a pattern of elevated Ab effector function emerged among subjects who experienced moderate or severe disease; those who experienced mild disease generated little activity.

In contrast to serum, but consistent with the lower relative levels of IgG Abs present in nasal wash samples, limited nasal ADNP, ADCC, and ADCD activity was observed (**Fig. 5b**). Comparison of serum and nasal effector functions showed positive correlation for ADCP activity, consistent with the observation that subjects with severe disease and high serum IgG also tended to exhibit high nasal IgG, and the known dependence of monocyte phagocytosis on IgG-binding FcγR (**Fig. 5c**).

### Ab features correlated with Ab functions

Next we established the characteristics of the Abs mediating each function by measuring the degree and direction of correlation between RBD-specific Ab biophysical features and Ab functions in serum (**Fig. 6a**). Consistent with their reliance on FcγR, ADCP, ADNP, and ADCC activities were most strongly correlated with FcγR binding, but also with levels of IgG1 and IgG3, which ligate FcγR best among human IgG subclasses. Interestingly, IgM positively correlated with both ADNP and neutralization activity, though the mechanistic relevance of each observed association is unclear. While IgA responses were robustly induced in serum, this isotype was generally more weakly associated with neutralization and effector activities.

**Figure 6.**
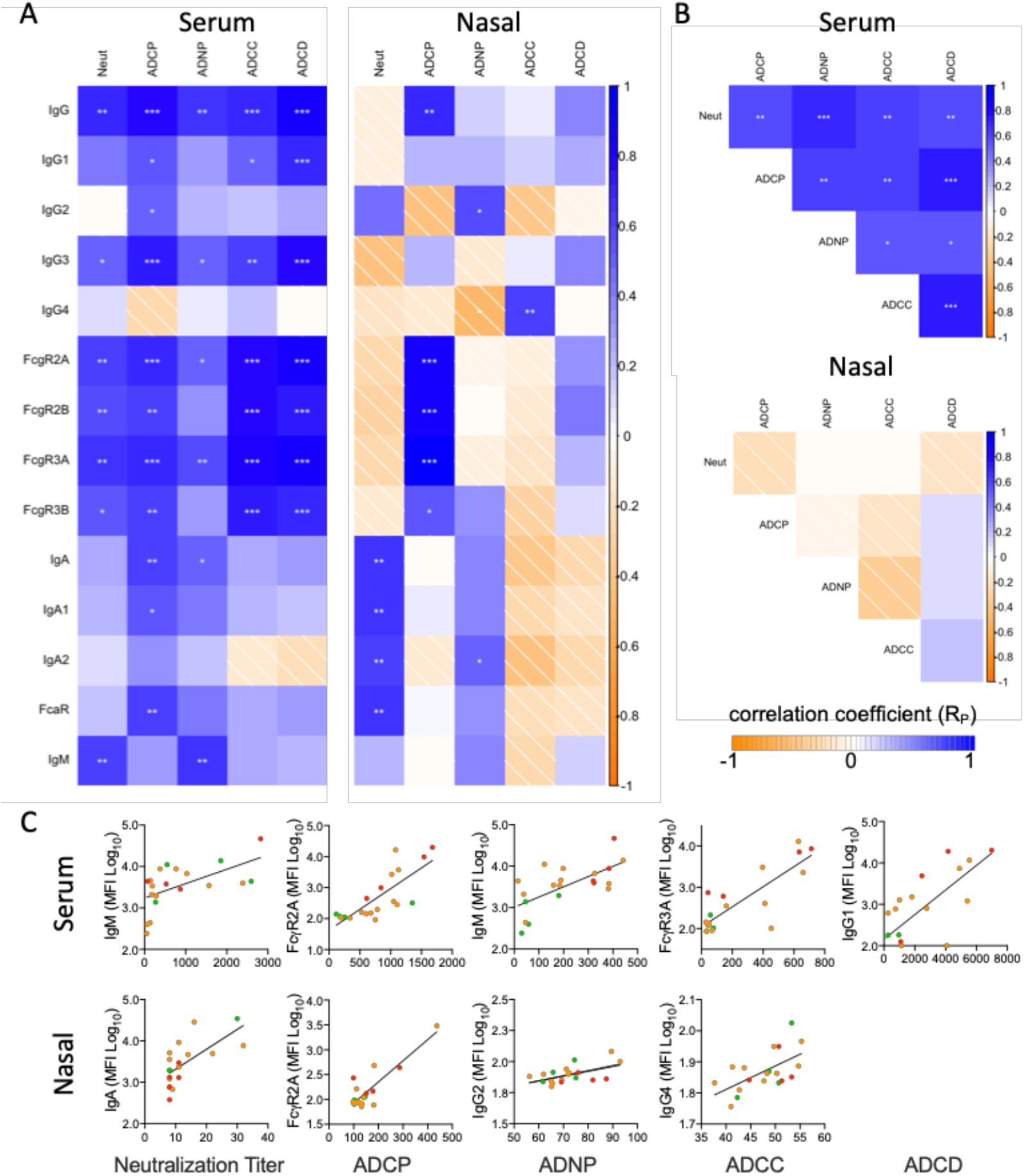
Correlative relationships between RBD-specific Ab features and functions. **A**. Correlations observed between RBD-specific Ab features and functions in serum (left) and nasal wash (right) samples. **B**. Correlations observed between Ab functions observed in serum (top) and nasal wash (bottom). **C**. Representative scatterplots between highly correlated Ab features and functions in serum (top) and nasal wash (bottom). *p < 0.05; **p < 0.01; ***p < 0.001. Pearson correlation coefficients (Rp) are shown.

In nasal samples, however, neutralization activity was strongly correlated with the IgA response (**Fig. 6a**. ADCP showed significant correlation with total RBD-specific IgG and FcγR-binding RBD-specific Abs. Strong relationships with the other effector functions, which generally showed low or negligible levels of activity, were generally not observed. Serum Ab functions were significantly correlated with one another; however, this relationship was not seen in nasal wash samples (**Fig. 6b**, consistent with the low activity observed for many functions, the differing isotypes associated with the robustly induced functions (neutralization and ADCP), and the bias observed among subjects to favor either a nasal IgA or a nasal IgG response. Representative scatterplots between individual features and functions show how these activities and characteristics varied by subject according to disease severity (**Fig. 6c**)

### Relationships between clinical characteristics and Ab responses

In order to probe how aspects of the immune response relate to subject characteristics, the magnitude and statistical confidence of differences between subjects by age, sex, and disease severity were evaluated (**Supplemental Fig. 11**). In exploratory analyses, comparisons were performed to determine which CoV-2-specific features showed differences between individuals experiencing severe versus mild or moderate disease in serum and nasal wash samples (unpaired two-sided test with Welch’s correction p < 0.05). As has been observed in other cohorts (Klein et al., 2020), a number of IgG-related responses were elevated in serum among individuals who had experienced severe disease (**Supplemental Fig. 12**). This elevation was also evident in nasal samples (**Fig. 7A**). Critically, RBD-specific Ab binding to FcαR was found to be significantly elevated in the nasal wash samples from subjects who had experienced mild or moderate as opposed to severe disease (**Fig. 7A**), and a number of other IgA-related responses exhibited differences near the arbitrary significance threshold (**Supplemental Fig. 11**). When examining the relationships between these features among donors who recovered from mild, moderate, and severe disease, IgG-related features typically showed a uniformly increasing magnitude with increasing disease severity (**Fig. 7B**). In contrast, the IgA-associated feature defined by nasal RBD-specific Abs binding to FcαR (**Fig. 7C**) and its correlated function neutralization (**Fig. 7D**) were lowest in subjects with either mild or severe disease, and elevated among those who recovered from moderate illness.

**Figure 7.**
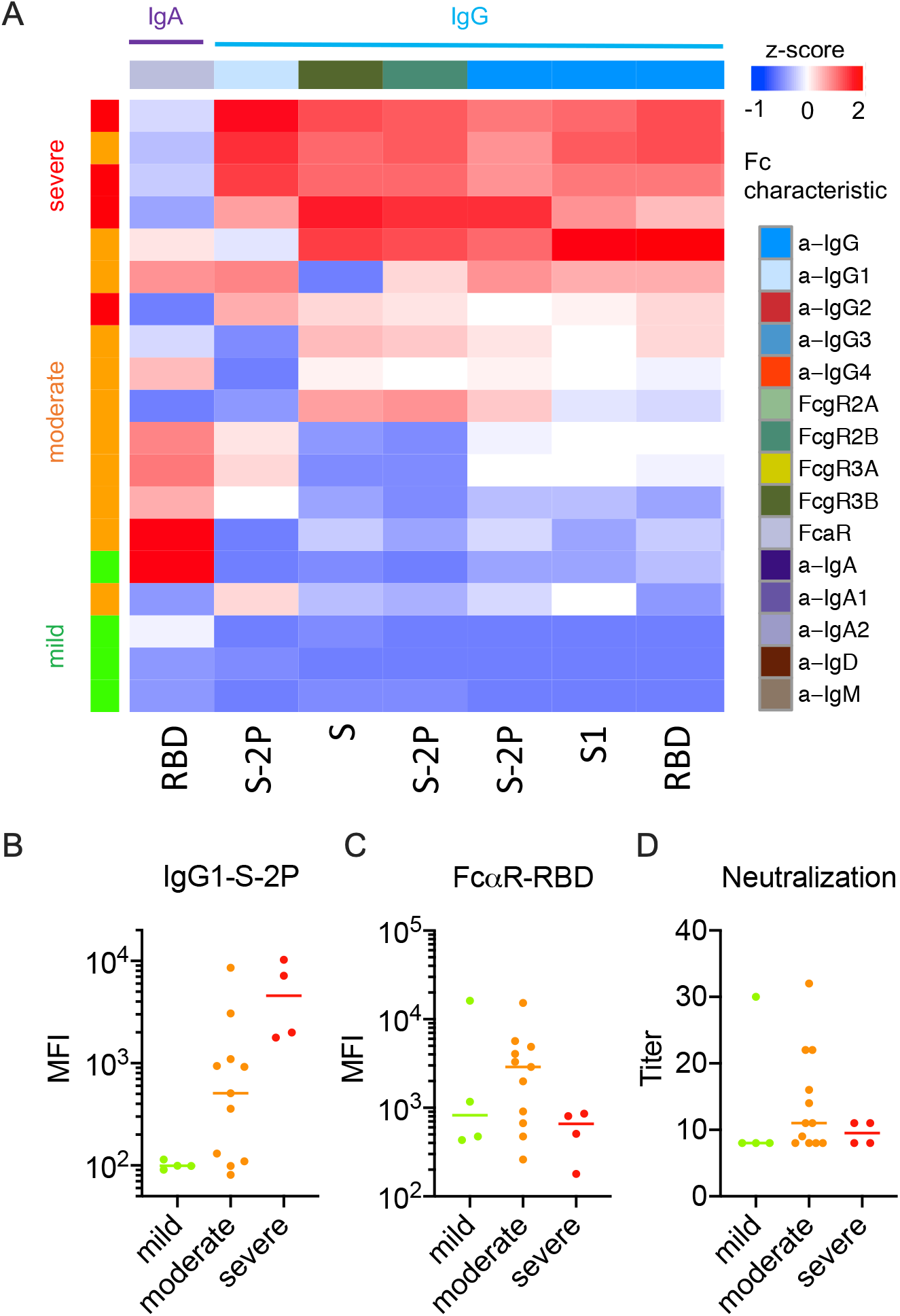
Nasal Ab features associated with severity. **A**. Heatmap of nasal CoV-2-specific Ab features that exhibited statistically significant differences in responses between subjects with severe and nonsevere disease (unpaired two-sided t-test with Welch’s correction, p < 0.05). **B-D**. Representative boxplots of nasal features by disease severity across donors who experienced mild, moderate, and severe disease. **B**. IgG1 specific to S-2P as a representative Ab feature elevated among subjects with severe disease. **C-D**. FcaR binding Abs specific to RBD (**C**), the sole Ab response elevated among subjects with non-severe disease, but which like nasal neutralization activity (**D**) is elevated among individuals who recovered from moderate disease as opposed to either mild or severe disease.

## Discussion

While information on systemic immune responses to SARS-CoV-2 continues to rapidly accrue, questions remain about mucosal immune responses to the virus within the respiratory tract - the primary site of SARS-CoV-2 infection and replication (Wölfel et al., 2020) and where IgA is the dominant Ab isotype (Chodirker and Tomasi, 1963; Conley and Delacroix, 1987; Tomasi et al., 1965). Prior work to delineate Ab responses to human CoV (Callow, 1985) and from studies of CoV strains in animals (Loa et al., 2002; Pearson et al., 2019; Saif, 1987), suggests that induction of mucosal Ab is a key component in reducing virus shedding after infection and may mediate protective immunity following re-exposure. Moreover, studies of mucosally-targeted SARS-CoV vaccines in animal models have identified virus-specific mucosal IgA as playing a role in protection from subsequent challenge (Du et al., 2008; See et al., 2006). Accordingly, strategies to prevent SARS-CoV-2 infection and disease are likely to benefit from not only robust systemic immunity, but also functional mucosal immunity to prevent infection at sites of virus entry (Moreno-Fierros et al., 2020). Indeed, robust mucosal immunity may serve not only to protect the individual, but also populations by reducing the level or duration of viral shedding.

We sought to characterize the humoral immune response against SARS-CoV-2 with an emphasis on Ab features and functions observed at distinct anatomical sites. We examined the CoV-specific Ab response across a panel of SARS-CoV-2 antigens, six other endemic human CoVs, and the putative precursor bat CoV, WIV1 (Zeng et al., 2016). Intriguingly, Abs that bound to endemic CoV also appeared to be boosted among SARS-CoV-2 convalescent donors. While the early appearance of CoV-specific IgG in a subset of patients (Long et al., 2020a; Zhao et al., 2020) is suggestive of a recall response, data presented here cannot define whether these boosted Abs are cross-reactive across CoV, or represent a more general boosting phenomenon. Consistent with the possibility that boosted Abs are cross-reactive, SARS-CoV-2 S-reactive CD4 T cells, a prerequisite for class-switched Ab responses, have been detected in the majority of COVID-19 patients, as well as in 34% of uninfected individuals, supporting the existence of shared epitopes between S proteins of endemic CoVs and SARS-CoV-2 (Braun et al., 2020; Grifoni et al., 2020a; Grifoni et al., 2020b). Additionally, immunofluorescence assays (Wölfel et al., 2020; Yuan et al., 2020) have exhibited a similar phenomenon, whereas Abs specific to the control antigens from other mucosal pathogens tested in this study were not elevated among convalescent donors. While many possibilities exist, the observation that the aged are most prone to severe COVID-19 illness, combined with the fact that the elderly have a decreased ability to generate *de novo* Abs (de Bourcy et al., 2017; Gibson et al., 2009; Henry et al., 2019), raises the possibility that pre-existing immunological memory resulting in induction of poorly neutralizing but cross-reactive Abs from prior exposure to circulating CoVs such as OC43, 229E, HKU1, and NL63 (Ng et al., 2020) may be associated with severe COVID-19 illness (Tetro, 2020).

Both multiplexed Ab profiling and functional assays showed induction of a generally robust humoral response to infection with SARS-CoV-2 in the majority of subjects, but an occasional lack of seroconversion in the context of mild disease (Klein et al., 2020; Sekine et al., 2020). Indeed, the magnitude of the humoral response in serum appeared to be closely tied to the clinical severity of infection. Closer dissection of the humoral response revealed that it was primarily comprised of IgG1 and IgG3 - subclasses that readily promote effector function via their Fc domains and which were, along with FcγR-binding SARS-CoV-2-specific Abs, associated with diverse effector functions. These observations suggest that SARS-CoV-2-specific Ab have the potential to contribute to protection against COVID-19 disease through the involvement of cells of the innate immune system and the complement system, and not solely by neutralization. While much has been made of the potential for Ab responses to promote infection or inflammation via interactions with FcR or via other mechanisms (Arvin et al., 2020; Eroshenko et al., 2020; Wang et al., 2016), it remains unclear whether the elevated Ab response magnitude is either a cause or effect of increased disease severity (Huang et al., 2020).

Separate from the dichotomy of whether Abs serve a protective or pathogenic role, we observed that the characteristics of the responses observed in serum and nasal samples tended to be highly distinct. Not only did the Ab profile of the nasal wash samples from individual subjects tend to favor either IgG or IgA to the exclusion of the other, there was an inverse correlation observed between the dominant nasal isotype and the prevalence of that isotype in serum. SARS-CoV-2-specific IgG in serum was associated with neutralization activity, consistent with prior work showing that neutralizing serum Abs against SARS-CoV-1 and SARS-CoV-2 are highly correlated with CoV-2-specific IgG (Cao et al., 2007; Klein et al., 2020). In contrast, those subjects who mounted a relatively IgA-biased nasal response exhibited elevated nasal wash neutralization activity, suggesting that the mechanistic contribution of mucosal IgA to neutralization of virus at the portal of entry could be substantial. Neutralization by mucosal IgA could be relevant *in vivo* in light of the observation that those subjects whose nasal specimens had the greatest neutralization potency also tended to report experiencing only mild or moderate symptoms. It is important to note however that the available cohort was not large enough to adequately power a robust examination of this trend. Reduced SARS-CoV-2-specific humoral responses among subjects with mild disease also confound the ability to identify potential correlates of protection among these convalescent donors. Further, because of the many correlations, inverse and direct, observed between Ab types and activities in different anatomical sites, depletion and other follow up studies will be needed to define the mechanistic relevance of feature-function correlations and their potential to make causal contributions to modifying disease severity. Nonetheless, taken together with prior studies of other CoV in humans and animals (Callow, 1985; Du et al., 2008; Loa et al., 2002; Pearson et al., 2019; Saif, 1987; See et al., 2006), these data raise the possibility that levels of SARS-CoV-2-specific mucosal IgA could serve as a useful immune correlate for mitigated disease severity, protection from infection, as well as reduced likelihood of transmission.

Chief among the limitations of this work is the small sample size, linked to low local disease prevalence. Other limitations include reliance on self-reported disease severity, and cross-sectional analysis of responses at a somewhat variable period after diagnosis and following varying degree and duration of symptoms among subjects with varying co-morbidities. Additionally, as with most mucosal sampling techniques, there was variability in the volume of nasal wash collected from each subject. This variability appeared to be largely due to accidental ingestion of the fluid during collection and affected volume to a greater degree than concentration. Experimental limitations include the use of lab-adapted cell lines rather than autologous or primary cells for evaluating some Ab functions. Surrogate endpoints of anti-viral activities, such as the substitution of FcγR3a activation and complement C3b deposition were employed as alternatives to assessing infected cell death or viral lysis, and compromises were made in antigenic fidelity by use of recombinant spike proteins. However, assays simplified in these ways may represent approaches with a broad ability to be deployed in global efforts to understand responses to infection and define protective immunity.

These compromises notwithstanding, these data have important implications for our understanding of the protection afforded by vaccination or prior infection. When considering vaccine development, an ideal candidate would not only protect the recipient from disease but would also prevent them from serving as an asymptomatic vector, as can be the case in other vaccine-treatable diseases such as polio and pertussis (Althouse and Scarpino, 2015; Holmgren and Czerkinsky, 2005; Warfel et al., 2014). Polio is a particularly informative model in this respect as the mucosally administered form of the vaccine is capable of providing sterilizing immunity - at the expense of a risk of reversion to virulence - while the systemically administered form fails to induce mucosal immunity and thus serves primarily to protect the recipient (Wright et al., 2016). Our current observation that natural infection elicits alternatively IgG or IgA-biased responses, with IgG associated with serum neutralization potency but severe disease and IgA associated with nasal neutralization activity and mild to moderate disease, suggests that this dichotomy could exist for COVID-19 as well. While human correlates of protection against SARS-CoV-2 in humans have yet to be defined, lessons from related CoV in animals and humans are consistent with the results of this small natural infection history study; mucosal IgA is likely of substantial importance.

## Methods

### Human subjects

A total of 35 individuals were studied, including 20 who had recovered from COVID-19 (age range: 18-77, mean: 53 yrs) and 15 naïve control subjects (age range: 22-66, mean: 40 yrs). Infection with SARS-CoV-2 was confirmed in all COVID-19 patients by real-time reverse-transcriptase-polymerase-chain-reaction of a nasopharyngeal swab. Study subjects included both males (17) and females (18). Disease severity among COVID-19 subjects ranged from mild (4) to moderate (12) and severe (4). Classification of disease severity was based on self-reported symptoms for individuals with mild or moderate disease, while a designation of severe disease was made on the basis of hospitalization for COVID-19. Serum, nasal wash, and stool samples were collected from each donor approximately one month after symptom onset, or one month from first positive PCR test in the case of asymptomatic (mild) disease (range: 19-67 days, mean: 40 days).

Primary neutrophils used in functional assays were purified from deidentified blood samples from healthy male and female donors over the age of 18 yrs. All research involving human subjects was approved by the Dartmouth College and Dartmouth-Hitchcock Medical Center Committee for the Protection of Human Subjects (Institutional Review Board) and written informed consent was obtained from all participants.

### Antigen and Fc Receptor expression and purification

Prefusion-stabilized, trimer-forming spike protomers (S-2P) of SARS-CoV-2, closely related and/or epidemic strains (SARS-CoV-1, WIV1, and MERS), and endemic coronaviruses (229E, OC43, NL63, and HKU1), and the receptor-binding domain of SARS-CoV-2 fused to a monomeric form of the human IgG4 Fc region were transiently expressed in either Expi 293 or Freestyle 293-F cells, and purified via affinity chromatography according to the manufacturers’ protocols (**Supplemental Tables 1 and 2**). Human FcγR were expressed and purified as described previously (Boesch et al., 2014).

### Fc array assay

CoV and control antigens, including S trimers, S subdomains *(i.e*., S1 and S2), and other viral proteins from SARS-CoV-2 as well as S and S subdomains from SARS CoV-1, MERS, HKU1, OC43, NL63, 229E, and WIV1 (**Supplemental Table 2**) and influenza HA and herpes simplex virus (HSV) gE proteins were covalently coupled to Luminex Magplex magnetic microspheres using a two-step carbodiimide chemistry as previously described (Brown et al., 2012). Biotinylated SARS-CoV-2 fusion peptide was captured on neutravidin-coupled microspheres. Pooled polyclonal serum IgG (IVIG), CR3022, a SARS CoV-1-specific monoclonal Ab that cross-reacts with SARS-CoV-2 S (Yuan et al., 2020), and VRC01, an HIV-specific monoclonal Ab, were used as controls to define the antigenicity profiles. The optimal dilution of serum was determined in pilot experiments in which a subset of samples was titrated. Test concentrations for serum ranged from 1:250 to 1:5000 and varied per detection reagent. Nasal wash and stool samples were assayed at a 1:10 dilution. Isotypes and subclasses of antigen-specific Abs were detected using R- phycoerthrin (PE) conjugated secondary Abs and by FcRs tetramers (**Supplemental Table 3**) as previously described (Brown et al., 2017; Brown et al., 2018). A FlexMap 3D array reader detected the beads and measured PE fluorescence used to calculate the Median Fluorescence Intensity (MFI).

### Neutralization assay

Samples of serum and nasal wash from SARS-CoV-2 convalescent and naïve donors were tested in microneutralization assays using a VSV-SARS-CoV pseudovirus system (Letko et al., 2020). In brief, samples were serially diluted 2-fold (1:50-1:3200 for serum; 1:4-1:256 for nasal wash) and incubated with a standardized concentration of SARS-CoV-2 pseudovirus for 1 hr at 37°C followed by addition to duplicate wells of 293T-ACE2-expressing target cells (Integral Molecular, Philadelphia PA) in a final volume of 100 ^.l per well. Plates were incubated at 37°C for 18-24 hrs, after which luciferase activity was measured using the Bright-Glo system (Promega, Madison WI) in a Bio-Tek II plate reader. Results were quantified relative to controls and data expressed as 60% neutralization titers.

### Phagocytosis assays

Assays of Ab-dependent phagocytosis by monocytes (ADCP) and neutrophils (ADNP) were performed essentially as described (Ackerman et al., 2011; Karsten et al., 2019; McAndrew et al., 2011). Briefly, 1 ^m yellow-green fluorescent microspheres (Thermo, F8813) were covalently conjugated with recombinant RBD and incubated for 3 hrs with dilute serum or nasal wash specimens and either the human monocytic THP-1 cell line (ATCC, TIB-202), or with freshly-isolated primary neutrophils. After pelleting, washing, and fixing, phagocytic scores were quantified as the product of the percentage of cells that phagocytosed one or more fluorescent beads and the median fluorescent intensity of this population as measured by flow cytometry with a MACSQuant Analyzer (Miltenyi Biotec). ADCP assays were performed in duplicate with high correspondence between results presented here and the replicate run. ADNP assays were performed in biological replicate using neutrophils purified from two different healthy donors for which results were averaged. A subset of neutrophils was stained with CD66b-APC (Biolegend G10F5) and PI (Biotium 41007) to determine the purity and viability of the isolated cellular fraction. CR3022 and VRC01 were used as positive and negative controls, respectively. Wells containing no Ab were used to define the level of Ab-independent phagocytosis.

### CD16 reporter assay

The ADCC potential of the specimens was measured using a Jurkat Lucia NFAT cell line (Invivogen, jktl-nfat-cd16), cultured according to the manufacturer’s recommendations, in which engagement of FcyR3a (CD16) on the cell surface leads to the secretion of luciferase. One day prior to running the assay, a high binding 96 well plate was coated with 1 ^g/mL SARS-CoV-2 RBD at 4°C overnight. Plates were then washed with PBS + 0.1% Tween20 and blocked at room temperature for 1 hr with PBS + 2.5% BSA. After washing, dilute serum or nasal wash sample and 100,000 cells/well in growth medium lacking antibiotics were cultured at 37°C for 24 hrs in a 200 ^l volume. The following day, 25 ^L of supernatant was drawn from each well and transferred to an opaque, white 96 well plate, to which 75 ^L of QuantiLuc substrate was added and luminescence immediately read on a SpectraMax Paradigm plate reader (Molecular Devices) using 1 s of integration time. The reported values are the mean of three kinetic reads taken at 0, 2.5, and 5 min. Negative control wells substituted assay medium for sample while 1x cell stimulation cocktail (Thermo, 00-4970-93) plus an additional 2 ^g/mL ionomycin were used to induce expression of the transgene as a positive control.

### Complement deposition assay

Antibody-dependent Complement Deposition (ADCD) was quantified essentially as previously described (Fischinger et al., 2019). In brief, serum and nasal samples were heat-inactivated at 56°C for 30 min prior to a 2 hr incubation at a dilution of 1:20 at RT with multiplex assay microspheres. After washing, each sample was incubated with Human Complement Serum (Sigma #S1764) at a concentration of 1:50 at RT with shaking for 1 hour. Samples were washed, sonicated, and incubated with murine anti-C3b (Cedarlane #CL7636AP) at RT for 1 hr followed by anti-mouse IgG1-PE secondary Ab (Southern Biotech #1070-09) at RT for 30 min. After a final wash and sonication, samples were resuspended in Luminex Shealth Fluid and complement deposition was determined on a MAGPIX (Luminex Corp) instrument to define the MFI. Assays performed without Ab and with heat-inactivated Human Complement Serum were used as negative controls.

### Data analysis and visualization

Basic analysis and visualization were performed using GraphPad Prism. Heatmaps, correlation plots, and boxplots were made in R (supported by R packages pheatmap, corrplot, and ggplot2). Hierarchical clustering was used to cluster and visualize data using the Manhattan and Euclidean metrics. Fc Array features were filtered by elimination of measurements for which >25% of the samples exhibited signal within 10 standard deviations (SD) of the technical blank. Fc Array features were log transformed, then scaled and centered by their standard deviation from the mean (z-score). A student’s two-tailed t-test with Welch’s correction with a cutoff of p= 0.05 was used to define features different between groups. Pearson correlation coefficients were calculated for the correlation matrices.

### Lead Contact

Further information and requests for resources and reagents should be directed to and will be fulfilled by the Lead Contact, Margaret E. Ackerman.

### Materials Availability

No new materials were created for this study.

### Data and Code Availability

Data and code to reproduce analyses are available at (link pending).

## Data Availability

Data will be made available upon request following peer review and publication in an archival journal.

## Acknowledgements

CoV S-2P and RBD-Fc expression constructs were provided by Dr. Jason McLellan (UT Austin), and fusion peptide was provided by Dr. Laura Walker and Mrunal Sakharkar (Adimab). The following reagent was produced under HHSN272201400008C and obtained through BEI Resources, NIAID, NIH: Spike Glycoprotein Receptor Binding Domain (RBD) from SARS-Related Coronavirus 2, Wuhan-Hu-1 with C-Terminal Histidine Tag, Recombinant from Baculovirus, NR-52307. This work was supported by NIH NCI supplement to 2 P30 CA 023108-41, the BioMT Molecular Tools Core supported by NIGMS COBRE award P20-GM113132, and by DHMC for providing support for collection of initial cohort samples. S.E.B. is supported by NIH NIAID 2T32AI007363. The authors thank Alejandra Prevost-Reilly, Dr. Anais Ovalle, and Dr. David de Gijsel for support of clinical specimen collection and donor enrollment, Drs. Paul and Cheryl Guyre for critical feedback and editorial assistance, and Dr. Paul Guyre, Jane Collins, and Jonell Hamilton for blood samples and assistance with neutrophil studies.

## Contributions

Conceptualization, P.F.W and M.E.A; Investigation, S.E.B., A.R.C., H.N., W.W.-A., and R.I.C.; Validation, S.X. and J.A.W.; Writing - original draft, S.E.B., A.R.C., H.N., J.L., M.E.A.; Writing - review and editing, all authors; Data Curation, S.X., J.A.W., R.I.C.; Supervision, P.F.W. and M.E.A.; Project Administration, J.A.W.; Funding acquisition, S.E.B., P.F.W, and M.E.A.

## References

Ackerman, M.E., Moldt, B., Wyatt, R.T., Dugast, A.S., McAndrew, E., Tsoukas, S., Jost, S., Berger, C. T., Sciaranghella, G., Liu, Q., et al. (2011). A robust, high-throughput assay to determine the phagocytic activity of clinical antibody samples. J Immunol Methods 366, 8–19.

Althouse, B.M., and Scarpino, S.V. (2015). Asymptomatic transmission and the resurgence of Bordetella pertussis. BMC Med 13, 146.

Arvin, A.M., Fink, K., Schmid, M.A., Cathcart, A., Spreafico, R., Havenar-Daughton, C., Lanzavecchia, A., Corti, D., and Virgin, H.W. (2020). A perspective on potential antibody-dependent enhancement of SARS-CoV-2. Nature.

Barouch, D.H., Alter, G., Broge, T., Linde, C., Ackerman, M.E., Brown, E.P., Borducchi, E.N., Smith, K.M., Nkolola, J.P., Liu, J., et al. (2015). Protective efficacy of adenovirus/protein vaccines against SIV challenges in rhesus monkeys. Science (New York, NY) 349, 320–324.

Boesch, A.W., Brown, E.P., Cheng, H.D., Ofori, M.O., Normandin, E., Nigrovic, P.A., Alter, G., and Ackerman, M.E. (2014). Highly parallel characterization of IgG Fc binding interactions. MAbs 6, 915–927.

Braun, J., Loyal, L., Frentsch, M., Wendisch, D., Georg, P., Kurth, F., Hippenstiel, S., Dingeldey, M., Kruse, B., Fauchere, F., et al. (2020). Presence of SARS-CoV-2 reactive T cells in COVID- 19 patients and healthy donors. medRxiv, 2020.2004.2017.20061440.

Brown, E.P., Dowell, K.G., Boesch, A.W., Normandin, E., Mahan, A.E., Chu, T., Barouch, D.H., Bailey-Kellogg, C., Alter, G., and Ackerman, M.E. (2017). Multiplexed Fc array for evaluation of antigen-specific antibody effector profiles. J Immunol Methods 443, 33–44.

Brown, E.P., Licht, A.F., Dugast, A.S., Choi, I., Bailey-Kellogg, C., Alter, G., and Ackerman, M.E. (2012). High-throughput, multiplexed IgG subclassing of antigen-specific antibodies from clinical samples. J Immunol Methods 386, 117–123.

Brown, E.P., Weiner, J.A., Lin, S., Natarajan, H., Normandin, E., Barouch, D.H., Alter, G., Sarzotti-Kelsoe, M., and Ackerman, M.E. (2018). Optimization and qualification of an Fc Array assay for assessments of antibodies against HIV-1/SIV. J Immunol Methods 455, 24–33.

Callow, K.A. (1985). Effect of specific humoral immunity and some non-specific factors on resistance of volunteers to respiratory coronavirus infection. J Hyg (Lond) 95, 173–189.

Cao, W.C., Liu, W., Zhang, P.H., Zhang, F., and Richardus, J.H. (2007). Disappearance of antibodies to SARS-associated coronavirus after recovery. N Engl J Med 357, 1162–1163.

Cao, Y., Su, B., Guo, X., Sun, W., Deng, Y., Bao, L., Zhu, Q., Zhang, X., Zheng, Y., Geng, C., et al. (2020). Potent neutralizing antibodies against SARS-CoV-2 identified by high-throughput single-cell sequencing of convalescent patients’ B cells. Cell.

Chan, C.M., Tse, H., Wong, S.S.Y., Woo, P.C.Y., Lau, S.K.P., Chen, L., Zheng, B.J., Huang, J.D., and Yuen, K.Y. (2009). Examination of seroprevalence of coronavirus HKU1 infection with S protein-based ELISA and neutralization assay against viral spike pseudotyped virus. J Clin Virol 45, 54–60.

Chandrashekar, A., Liu, J., Martinot, A.J., McMahan, K., Mercado, N.B., Peter, L., Tostanoski, L.H., Yu, J., Maliga, Z., Nekorchuk, M., et al. (2020). SARS-CoV-2 infection protects against rechallenge in rhesus macaques. Science (New York, NY).

Chen, X., Li, R., Pan, Z., Qian, C., Yang, Y., You, R., Zhao, J., Liu, P., Gao, L., Li, Z., et al. (2020). Human monoclonal antibodies block the binding of SARS-CoV-2 spike protein to angiotensin converting enzyme 2 receptor. Cellular & Molecular Immunology.

Chodirker, W.B., and Tomasi, T.B., Jr. (1963). Gamma-Globulins: Quantitative Relationships in Human Serum and Nonvascular Fluids. Science (New York, NY) 142, 1080–1081.

Cong, Y., Hart, B.J., Gross, R., Zhou, H., Frieman, M., Bollinger, L., Wada, J., Hensley, L.E., Jahrling, P.B., Dyall, J., et al. (2018). MERS-CoV pathogenesis and antiviral efficacy of licensed drugs in human monocyte-derived antigen-presenting cells. PLoS One 13, e0194868.

Conley, M.E., and Delacroix, D.L. (1987). Intravascular and mucosal immunoglobulin A: two separate but related systems of immune defense? Ann Intern Med 106, 892–899.

de Bourcy, C.F., Angel, C.J., Vollmers, C., Dekker, C.L., Davis, M.M., and Quake, S.R. (2017). Phylogenetic analysis of the human antibody repertoire reveals quantitative signatures of immune senescence and aging. Proc Natl Acad Sci U S A 114, 1105–1110.

Du, L., Zhao, G., Lin, Y., Sui, H., Chan, C., Ma, S., He, Y., Jiang, S., Wu, C., Yuen, K.Y., et al. (2008). Intranasal vaccination of recombinant adeno-associated virus encoding receptor-binding domain of severe acute respiratory syndrome coronavirus (SARS-CoV) spike protein induces strong mucosal immune responses and provides long-term protection against SARS-CoV infection. J Immunol 180, 948–956.

Duan, K., Liu, B., Li, C., Zhang, H., Yu, T., Qu, J., Zhou, M., Chen, L., Meng, S., Hu, Y., et al. (2020). Effectiveness of convalescent plasma therapy in severe COVID-19 patients. Proc Natl Acad Sci U S A 117, 9490–9496.

Eroshenko, N., Gill, T., Keaveney, M.K., Church, G.M., Trevejo, J.M., and Rajaniemi, H. (2020). Implications of antibody-dependent enhancement of infection for SARS-CoV-2 countermeasures. Nat Biotechnol 38, 789–791.

Fischinger, S., Fallon, J.K., Michell, A.R., Broge, T., Suscovich, T.J., Streeck, H., and Alter, G. (2019). A high-throughput, bead-based, antigen-specific assay to assess the ability of antibodies to induce complement activation. J Immunol Methods 473, 112630.

Gallais, F., Velay, A., Wendling, M.-J., Nazon, C., Partisani, M., Sibilia, J., Candon, S., and Fafi-Kremer, S. Intrafamilial Exposure to SARS-CoV-2 Induces Cellular Immune Response without Seroconversion. medRxiv.

Gibson, K.L., Wu, Y.C., Barnett, Y., Duggan, O., Vaughan, R., Kondeatis, E., Nilsson, B.O., Wikby, A., Kipling, D., and Dunn-Walters, D.K. (2009). B-cell diversity decreases in old age and is correlated with poor health status. Aging Cell 8, 18–25.

Grifoni, A., Sidney, J., Zhang, Y., Scheuermann, R.H., Peters, B., and Sette, A. (2020a). A Sequence Homology and Bioinformatic Approach Can Predict Candidate Targets for Immune Responses to SARS-CoV-2. Cell Host & Microbe 27, 671–680.e672.

Grifoni, A., Weiskopf, D., Ramirez, S.I., Mateus, J., Dan, J.M., Moderbacher, C.R., Rawlings, S.A., Sutherland, A., Premkumar, L., Jadi, R.S., et al. (2020b). Targets of T Cell Responses to SARS-CoV-2 Coronavirus in Humans with COVID-19 Disease and Unexposed Individuals. Cell 181, 1489–1501 e1415.

Guan, W.J., Ni, Z.Y., Hu, Y., Liang, W.H., Ou, C.Q., He, J.X., Liu, L., Shan, H., Lei, C.L., Hui, D.S.C., et al. (2020). Clinical Characteristics of Coronavirus Disease 2019 in China. N Engl J Med 382, 1708–1720.

Hassan, A.O., Case, J.B., Winkler, E.S., Thackray, L.B., Kafai, N.M., Bailey, A.L., McCune, B.T., Fox, J.M., Chen, R.E., Alsoussi, W.B., et al. (2020). A SARS-CoV-2 Infection Model in Mice Demonstrates Protection by Neutralizing Antibodies. Cell.

Henry, C., Zheng, N.Y., Huang, M., Cabanov, A., Rojas, K.T., Kaur, K., Andrews, S.F., Palm, A.E., Chen, Y.Q., Li, Y., et al. (2019). Influenza Virus Vaccination Elicits Poorly Adapted B Cell Responses in Elderly Individuals. Cell Host Microbe 25, 357–366 e356.

Hoeppel, W., Chen, H.-J., Allahverdiyeva, S., Manz, X., Aman, J., Biobank, A.U.C.-., Bonta, P., Brouwer, P., de Taeye, S., Caniels, T., et al. (2020). Anti-SARS-CoV-2 IgG from severely ill COVID-19 patients promotes macrophage hyper-inflammatory responses.

Holmgren, J., and Czerkinsky, C. (2005). Mucosal immunity and vaccines. Nat Med 11, S45-53.

Huang, A.T., Garcia-Carreras, B., Hitchings, M.D.T., Yang, B., Katzelnick, L.C., Rattigan, S.M., Borgert, B.A., Moreno, C.A., Solomon, B.D., Rodriguez-Barraquer, I., et al. (2020). A systematic review of antibody mediated immunity to coronaviruses: antibody kinetics, correlates of protection, and association of antibody responses with severity of disease. medRxiv.

Imai, M., Iwatsuki-Horimoto, K., Hatta, M., Loeber, S., Halfmann, P.J., Nakajima, N., Watanabe, T., Ujie, M., Takahashi, K., Ito, M., et al. (2020). Syrian hamsters as a small animal model for SARS-CoV-2 infection and countermeasure development. Proc Natl Acad Sci U S A 117, 16587–16595.

Jaume, M., Yip, M.S., Cheung, C.Y., Leung, H.L., Li, P.H., Kien, F., Dutry, I., Callendret, B., Escriou, N., Altmeyer, R., et al. (2011). Anti-severe acute respiratory syndrome coronavirus spike antibodies trigger infection of human immune cells via a pH- and cysteine protease- independent FcgammaR pathway. J Virol 85, 10582–10597.

Ju, B., Zhang, Q., Ge, J., Wang, R., Sun, J., Ge, X., Yu, J., Shan, S., Zhou, B., Song, S., et al. (2020). Human neutralizing antibodies elicited by SARS-CoV-2 infection. Nature.

Karsten, C.B., Mehta, N., Shin, S.A., Diefenbach, T.J., Slein, M.D., Karpinski, W., Irvine, E.B., Broge, T., Suscovich, T.J., and Alter, G. (2019). A versatile high-throughput assay to characterize antibody-mediated neutrophil phagocytosis. J Immunol Methods 471, 46–56.

Klein, S., Pekosz, A., Park, H.S., Ursin, R., Shapiro, J., Benner, S., Littlefield, K., Kumar, S., Naik, H.M., Betenbaugh, M., et al. (2020). Sex, age, and hospitalization drive antibody responses in a COVID-19 convalescent plasma donor population. medRxiv.

Le Bert, N., Tan, A.T., Kunasegaran, K., Tham, C.Y.L., Hafezi, M., Chia, A., Chng, M.H.Y., Lin, M., Tan, N., Linster, M., et al. (2020). SARS-CoV-2-specific T cell immunity in cases of COVID-19 and SARS, and uninfected controls. Nature.

Letko, M., Marzi, A., and Munster, V. (2020). Functional assessment of cell entry and receptor usage for SARS-CoV-2 and other lineage B betacoronaviruses. Nat Microbiol 5, 562–569.

Liu, L., Wei, Q., Lin, Q., Fang, J., Wang, H., Kwok, H., Tang, H., Nishiura, K., Peng, J., Tan, Z., et al. (2019). Anti-spike IgG causes severe acute lung injury by skewing macrophage responses during acute SARS-CoV infection. JCI Insight 4.

Loa, C.C., Lin, T.L., Wu, C.C., Bryan, T., Hooper, T., and Schrader, D. (2002). Specific mucosal IgA immunity in turkey poults infected with turkey coronavirus. Vet Immunol Immunopathol 88, 57–64.

Long, Q.-x., Deng, H.-j., Chen, J., Hu, J., Liu, B.-z., Liao, P., Lin, Y., Yu, L.-h., Mo, Z., Xu, Y.-y., et al. (2020a). Antibody responses to SARS-CoV-2 in COVID-19 patients: the perspective application of serological tests in clinical practice. medRxiv, 2020.2003.2018.20038018.

Long, Q.X., Tang, X.J., Shi, Q.L., Li, Q., Deng, H.J., Yuan, J., Hu, J.L., Xu, W., Zhang, Y., Lv, F.J., et al. (2020b). Clinical and immunological assessment of asymptomatic SARS-CoV-2 infections. Nat Med.

McAndrew, E.G., Dugast, A.S., Licht, A.F., Eusebio, J.R., Alter, G., and Ackerman, M.E. (2011). Determining the phagocytic activity of clinical antibody samples. J Vis Exp, e3588.

Moreno-Fierros, L., Garcia-Silva, I., and Rosales-Mendoza, S. (2020). Development of SARS-CoV- 2 vaccines: should we focus on mucosal immunity? Expert Opin Biol Ther.

Ng, K., Faulkner, N., Cornish, G., Rosa, A., Earl, C., Wrobel, A., Benton, D., Roustan, C., Bolland, W., Thompson, R., et al. (2020). Pre-existing and de novo humoral immunity to SARS-CoV-2 in humans. bioRxiv, 2020.2005.2014.095414.

Pearson, M., LaVoy, A., Evans, S., Vilander, A., Webb, C., Graham, B., Musselman, E., LeCureux, J., VandeWoude, S., and Dean, G.A. (2019). Mucosal Immune Response to Feline Enteric Coronavirus Infection. Viruses 11.

Pinto, D., Park, Y.J., Beltramello, M., Walls, A.C., Tortorici, M.A., Bianchi, S., Jaconi, S., Culap, K., Zatta, F., De Marco, A., et al. (2020). Cross-neutralization of SARS-CoV-2 by a human monoclonal SARS-CoV antibody. Nature.

Pittala, S., Morrison, K.S., and Ackerman, M.E. (2019). Systems serology for decoding infection and vaccine-induced antibody responses to HIV-1. Curr Opin HIV AIDS 14, 253–264.

Rogers, T.F., Zhao, F., Huang, D., Beutler, N., Burns, A., He, W.T., Limbo, O., Smith, C., Song, G., Woehl, J., et al. (2020). Isolation of potent SARS-CoV-2 neutralizing antibodies and protection from disease in a small animal model. Science (New York, NY).

Saif, L.J. (1987). Development of nasal, fecal and serum isotype-specific antibodies in calves challenged with bovine coronavirus or rotavirus. Vet Immunol Immunopathol 17, 425–439.

Salazar, E., Perez, K.K., Ashraf, M., Chen, J., Castillo, B., Christensen, P.A., Eubank, T., Bernard, D. W., Eagar, T.N., Long, S.W., et al. (2020). Treatment of Coronavirus Disease 2019 (COVID- 19) Patients with Convalescent Plasma. The American journal of pathology.

See, R.H., Zakhartchouk, A.N., Petric, M., Lawrence, D.J., Mok, C.P., Hogan, R.J., Rowe, T., Zitzow, L.A., Karunakaran, K.P., Hitt, M.M., et al. (2006). Comparative evaluation of two severe acute respiratory syndrome (SARS) vaccine candidates in mice challenged with SARS coronavirus. J Gen Virol 87, 641–650.

Sekine, T., Perez-Potti, A., Rivera-Ballesteros, O., Strålin, K., Gorin, J.-B., Olsson, A., Llewellyn-Lacey, S., Kamal, H., Bogdanovic, G., Muschiol, S., et al. (2020). Robust T cell immunity in convalescent individuals with asymptomatic or mild COVID-19. bioRxiv, 2020.2006.2029.174888.

Seow, J., Graham, C., Merrick, B., Acors, S., Steel, K.J.A., Hemmings, O., A., O.B., Kouphou, N., Pickering, S., Galao, R., et al. Longitudinal evaluation and decline of antibody responses in SARS-CoV-2 infection. medRxiv.

Shen, C., Wang, Z., Zhao, F., Yang, Y., Li, J., Yuan, J., Wang, F., Li, D., Yang, M., Xing, L., et al. (2020). Treatment of 5 Critically Ill Patients With COVID-19 With Convalescent Plasma. JAMA.

Shi, R., Shan, C., Duan, X., Chen, Z., Liu, P., Song, J., Song, T., Bi, X., Han, C., Wu, L., et al. (2020). A human neutralizing antibody targets the receptor binding site of SARS-CoV-2. Nature.

Sui, J., Li, W., Murakami, A., Tamin, A., Matthews, L.J., Wong, S.K., Moore, M.J., Tallarico, A.S.C., Olurinde, M., Choe, H., et al. (2004). Potent neutralization of severe acute respiratory syndrome (SARS) coronavirus by a human mAb to S1 protein that blocks receptor association. Proceedings of the National Academy of Sciences of the United States of America 101, 2536.

ter Meulen, J., van den Brink, E.N., Poon, L.L.M., Marissen, W.E., Leung, C.S.W., Cox, F., Cheung, C.Y., Bakker, A.Q., Bogaards, J.A., van Deventer, E., et al. (2006). Human Monoclonal Antibody Combination against SARS Coronavirus: Synergy and Coverage of Escape Mutants. PLOS Medicine 3, e237.

Tetro, J.A. (2020). Is COVID-19 receiving ADE from other coronaviruses? Microbes Infect 22, 72-73.

Tomasi, T.B., Jr., Tan, E.M., Solomon, A., and Prendergast, R.A. (1965). Characteristics of an Immune System Common to Certain External Secretions. J Exp Med 121, 101–124.

Traggiai, E., Becker, S., Subbarao, K., Kolesnikova, L., Uematsu, Y., Gismondo, M.R., Murphy, B. R., Rappuoli, R., and Lanzavecchia, A. (2004). An efficient method to make human monoclonal antibodies from memory B cells: potent neutralization of SARS coronavirus. Nature Medicine 10, 871–875.

Uddin, M.B., Hasan, M., Harun-Al-Rashid, A., Ahsan, M.I., Imran, M.A.S., and Ahmed, S.S.U. (2020). Ancestral origin, antigenic resemblance and epidemiological insights of novel coronavirus (SARS-CoV-2): Global burden and Bangladesh perspective. Infect Genet Evol 84, 104440.

Vennema, H., de Groot, R.J., Harbour, D.A., Dalderup, M., Gruffydd-Jones, T., Horzinek, M.C., and Spaan, W.J. (1990). Early death after feline infectious peritonitis virus challenge due to recombinant vaccinia virus immunization. J Virol 64, 1407–1409.

Wang, C., Li, W., Drabek, D., Okba, N.M.A., van Haperen, R., Osterhaus, A.D.M.E., van Kuppeveld, F.J.M., Haagmans, B.L., Grosveld, F., and Bosch, B.-J. (2020). A human monoclonal antibody blocking SARS-CoV-2 infection. bioRxiv, 2020.2003.2011.987958.

Wang, Q., Zhang, L., Kuwahara, K., Li, L., Liu, Z., Li, T., Zhu, H., Liu, J., Xu, Y., Xie, J., et al. (2016). Immunodominant SARS Coronavirus Epitopes in Humans Elicited both Enhancing and Neutralizing Effects on Infection in Non-human Primates. ACS Infect Dis 2, 361–376.

Warfel, J.M., Zimmerman, L.I., and Merkel, T.J. (2014). Acellular pertussis vaccines protect against disease but fail to prevent infection and transmission in a nonhuman primate model. Proc Natl Acad Sci U S A 111, 787–792.

Wec, A.Z., Wrapp, D., Herbert, A.S., Maurer, D.P., Haslwanter, D., Sakharkar, M., Jangra, R.K., Dieterle, M.E., Lilov, A., Huang, D., et al. (2020). Broad neutralization of SARS-related viruses by human monoclonal antibodies. Science (New York, NY).

Wölfel, R., Corman, V.M., Guggemos, W., Seilmaier, M., Zange, S., Müller, M.A., Niemeyer, D., Jones, T.C., Vollmar, P., Rothe, C., et al. (2020). Virological assessment of hospitalized patients with COVID-2019. Nature 581, 465–469.

Wright, P.F., Connor, R.I., Wieland-Alter, W.F., Hoen, A.G., Boesch, A.W., Ackerman, M.E., Oberste, M.S., Gast, C., Brickley, E.B., Asturias, E.J., et al. (2016). Vaccine-induced mucosal immunity to poliovirus: analysis of cohorts from an open-label, randomised controlled trial in Latin American infants. Lancet Infect Dis 16, 1377–1384.

Wu, Y., Wang, F., Shen, C., Peng, W., Li, D., Zhao, C., Li, Z., Li, S., Bi, Y., Yang, Y., et al. (2020). A noncompeting pair of human neutralizing antibodies block COVID-19 virus binding to its receptor ACE2. Science (New York, NY) 368, 1274–1278.

Xu, T.M., Lin, B., Chen, C., Liu, L.G., and Xue, Y. (2020). Non-optimal effectiveness of convalescent plasma transfusion and hydroxychloroquine in treating COVID-19: a case report. Virol J 17, 80.

Yang, Z.Y., Werner, H.C., Kong, W.P., Leung, K., Traggiai, E., Lanzavecchia, A., and Nabel, G.J. (2005). Evasion of antibody neutralization in emerging severe acute respiratory syndrome coronaviruses. Proc Natl Acad Sci U S A 102, 797–801.

Ye, L., Chen, X., Li, R., Pan, Z., Qian, C., Yang, Y., You, R., Zhao, J., Gao, L., Li, Z., et al. (2020). Human monoclonal antibodies block the binding of SARS-CoV-2 spike protein to angiotensin converting enzyme 2 receptor. medRxiv, 2020.2004.2006.20055475.

Yip, M.S., Leung, N.H., Cheung, C.Y., Li, P.H., Lee, H.H., Daeron, M., Peiris, J.S., Bruzzone, R., and Jaume, M. (2014). Antibody-dependent infection of human macrophages by severe acute respiratory syndrome coronavirus. Virol J 11, 82.

Yu, J., Tostanoski, L.H., Peter, L., Mercado, N.B., McMahan, K., Mahrokhian, S.H., Nkolola, J.P., Liu, J., Li, Z., Chandrashekar, A., et al. (2020). DNA vaccine protection against SARS-CoV-2 in rhesus macaques. Science (New York, NY).

Yuan, M., Liu, H., Wu, N.C., Lee, C.-C.D., Zhu, X., Zhao, F., Huang, D., Yu, W., Hua, Y., Tien, H., et al. (2020). Structural basis of a public antibody response to SARS-CoV-2. bioRxiv: the preprint server for biology, 2020.2006.2008.141267.

Zeng, L.P., Gao, Y.T., Ge, X.Y., Zhang, Q., Peng, C., Yang, X.L., Tan, B., Chen, J., Chmura, A.A., Daszak, P., et al. (2016). Bat Severe Acute Respiratory Syndrome-Like Coronavirus WIV1 Encodes an Extra Accessory Protein, ORFX, Involved in Modulation of the Host Immune Response. J Virol 90, 6573–6582.

Zeng, Q.L., Yu, Z.J., Gou, J.J., Li, G.M., Ma, S.H., Zhang, G.F., Xu, J.H., Lin, W.B., Cui, G.L., Zhang, M.M., et al. (2020). Effect of Convalescent Plasma Therapy on Viral Shedding and Survival in Patients With Coronavirus Disease 2019. J Infect Dis 222, 38–43.

Zhang, B., Liu, S., Tan, T., Huang, W., Dong, Y., Chen, L., Chen, Q., Zhang, L., Zhong, Q., Zhang, X., et al. (2020). Treatment With Convalescent Plasma for Critically Ill Patients With SARS-CoV- 2 Infection. Chest.

Zhao, J., Yuan, Q., Wang, H., Liu, W., Liao, X., Su, Y., Wang, X., Yuan, J., Li, T., Li, J., et al. (2020). Antibody responses to SARS-CoV-2 in patients of novel coronavirus disease 2019. Clin Infect Dis.

Zhu, Z., Chakraborti, S., He, Y., Roberts, A., Sheahan, T., Xiao, X., Hensley, L.E., Prabakaran, P., Rockx, B., Sidorov, I.A., et al. (2007). Potent cross-reactive neutralization of SARS coronavirus isolates by human monoclonal antibodies. Proceedings of the National Academy of Sciences 104, 12123.

